# GLLaucoMed: A Secure LLM-Powered Agentic Workflow for Automated Medication Extraction from Free-Text Glaucoma Clinical Notes

**DOI:** 10.64898/2026.06.12.26355525

**Authors:** Nicholas Solages, Rafael Scherer, Gustavo A. Samico, Naomi E. Gutkind, Janet Kang, Felipe A. Medeiros, Swarup S. Swaminathan

## Abstract

**Purpose:** To evaluate the efficacy of large language models (LLMs) in extracting medication-related information from glaucoma clinical notes in the electronic health record (EHR).

**Design:** Cross-sectional.

**Subjects:** 1,250 subjects in the Bascom Palmer Ophthalmic Repository.

**Methods:** Extracted clinical notes from glaucoma-related encounters between 2014 and 2024 were labeled by two glaucoma specialists with a third serving as an adjudicator. Graders were asked to label current topical medications (CTM), proposed changes to topical medications (ΔTM), current oral medications (COM), and proposed changes to oral medications (ΔOM) in a structured fashion. The dataset was split into development (10%), validation (10%), and test (80%) sets stratified by clinician. Development and validation sets were used to engineer and refine prompts, and the held-out test set was used for model assessment. Five LLMs (Claude Opus 4.6, DeepSeek-V3.2, GPT 5.2, Grok 4.1, and Qwen3.6-35B-A3B) were accessed via Microsoft Azure AI Foundry within a HIPAA-compliant environment. Inter-grader agreement was assessed with Gwet AC1. LLM performance was initially assessed in a binary fashion with F1 scores, and the degree of text match among positive cases was evaluated using exact match accuracy and Jaccard Index (JI).

**Main Outcome Measures:** F1 score, exact match accuracy, JI.

**Results:** Gwet AC1 for intergrader agreement was 0.799, 0.888, 0.985, and 0.988 for CTM, ΔTM, COM, and ΔOM, respectively. F1 scores for CTM were 0.985, 0.971, 0.978, 0.968, and 0.970 for Claude, Deepseek, GPT, Grok, and Qwen, respectively; for ΔTM: 0.905, 0.826, 0.897, 0.842, 0.855, respectively; for COM: 0.923, 0.887, 0.899, 0.906, 0.894, respectively; for ΔOM: 0.958, 0.815, 0.937, 0.835, 0.940, respectively. Among positive cases, range of exact match accuracies for CTM (N=1354) was 0.730-0.882 and range of JIs was 0.809-0.918. For ΔTM (N=404), exact match accuracy range was 0.619-0.780 and JI range was 0.668-0.827. For COM (N=47), exact match accuracy range was 0.766-0.872 and JI range was 0.765-0.870. For ΔOM (N=25), exact match accuracy range was 0.583-0.920 and JI range was 0.583-0.922.

**Conclusions:** The GLLaucoMed pipeline demonstrated high performance in extracting and standardizing medication data from unstructured clinical notes, including both current medications and proposed changes. Claude and GPT exhibited the strongest performance.

## Introduction

Glaucoma is the leading cause of irreversible blindness worldwide, affecting approximately 80 million people globally and 4 million Americans.^1, 2^ A chronic and irreversible disease, glaucoma is characterized by progressive optic nerve degeneration, with elevated intraocular pressure (IOP) as the only modifiable risk factor. Current glaucoma treatments include medications, laser therapies, and surgical interventions to reduce IOP.^3^ Given glaucoma is a lifelong condition, patients frequently require adjustments to their medication regimens, underscoring the importance of accurate medication assessments for both clinical care and research.

Electronic health records (EHRs) are now widely used in the United States,^4^ creating large databases that are being culled for research.^5^ “Big data” repositories including the American Academy of Ophthalmology Intelligent Research in Sight (IRIS) Registry and the National Institutes of Health All of Us program aggregate multicenter structured EHR data for research purposes. Such repositories are currently limited to structured outputs (i.e., standardized and organized data fields with limited and discrete values). Unstructured data such as free-text clinical notes are excluded, primarily due to potential data security concerns regarding protected health information (PHI).^6^ Clinical notes often contain details not captured by structured data fields; only 13% of concepts extracted from patient notes have similar structured counterparts,^7^ highlighting the substantial body of clinically relevant information that remains inaccessible when analyses are confined to structured fields. Free-text clinical notes represent a major opportunity for advancing research quality and depth.

Any research evaluating the efficacy of treatment requires a complete assessment of all available data. In glaucoma, the challenge of capturing accurate medication data from the EHR is particularly acute. Medication regimens of glaucoma patients are dynamic, changing frequently due to adverse effects, inefficacy, or other interventions. Such changes are not always accurately reflected in the EHR medication list. A manual chart review of 150 glaucoma outpatient visits found discrepancies in ophthalmic medications documented in medication lists and progress notes for 32% of medications.^8^ Similarly, Bacon et al. found that 20% of medication reconciliation records listed medications with instructions that were incorrect based on ophthalmologists’ treatment plans, and that when discrepancies existed, approximately 75% of patients followed the clinician-stated regimen rather than the EHR medication list.^9^ These findings demonstrate that structured medication lists in the EHR are frequently unreliable for glaucoma patients, and that the clinical note often represents the most accurate record. In large cohort studies, use of IOP data from the EHR without accounting for the degree of treatment can confound the relationship between IOP and glaucoma progression over time. Prior work has demonstrated that glaucoma outcomes can differ among eyes with similar IOP values but different treatment intensities due to diurnal IOP fluctuations and challenges with medication adherence.^10, 11^

Large language models (LLMs) are large neural networks trained on large bodies of data and are powerful tools capable of processing and extracting information from free-text sources. While earlier natural language processing (NLP) approaches relied on rule-based algorithms or traditional machine learning, the advent of LLMs has dramatically expanded the range and accuracy of information that can be automatically extracted. In ophthalmology, prior work demonstrated that medication entities such as drug name and frequency could be extracted from glaucoma progress notes with high accuracy.^12, 13^ However, these works used smaller datasets, did not evaluate labels at the note level, and did not evaluate documented changes in medication regimens. Prior work also did not utilize novel approaches such as agentic workflows. As an extension of LLMs, agentic workflows involved staged questioning with sequential reasoning, conditional branching, self-correction, and often autonomous decision-making. Agentic workflows are of significant interest in healthcare applications, both in the research and clinical domains.^14^

The purpose of this study was to compare the ability of leading LLMs to extract structured medication information from a large corpus of labeled free-text EHR clinical notes of glaucoma patients in a secure environment. We sought to utilize agentic workflows to extract not only the medications currently in use at the time of a clinical encounter, but also any proposed changes documented by the treating clinician within the progress note for both topical and oral medications.

## Methods

### Data Collection and Note Verification

This study was approved by the Institutional Review Board at the University of Miami. The requirement for informed consent was waived because of the retrospective nature of this work. This work adhered to the Declaration of Helsinki and complied with the Health Insurance Portability and Accountability Act (HIPAA) for maintaining patient confidentiality.

This study was a retrospective analysis of patients from the Bascom Palmer Ophthalmic Repository (BPOR), which contains data from over 70,000 patients evaluated at the Bascom Palmer Eye Institute (BPEI). This database contains demographic, exam, imaging, notes, and procedural data of eyes with glaucoma or suspected of having glaucoma examined at the clinical sites of the BPEI, identified using ICD codes from the EHR system (Epic Systems, Verona, WI). Prior work reviews the data extraction process in detail.^15, 16^ Only glaucoma-related encounters at BPEI were evaluated in this study.

Our prior work details the extraction of BPOR clinical notes.^17^ Briefly, we extracted clinical notes recorded between 2014 and 2024 from outpatient encounters with signed notes and glaucoma-related ICD codes (H40.X) listed as visit diagnoses. A total of 20 BPEI glaucoma specialists were represented in this data extract. All notes were stored on a HIPAA-compliant university-approved institutional server (Box, Redwood City, CA). We filtered notes to include only clinically relevant appointment types (e.g., new patient or follow-up visits). To ensure adequate data quality, we excluded notes with fewer than 200 characters. The dataset contained 1,250 notes corresponding to 1,250 unique subjects (one encounter note per patient). Of these, we discovered eight notes to be attestations by supervising physicians rather than clinical notes, which were excluded from subsequent analysis, leaving a total of 1,242 notes in the final dataset.

### Note Labeling

Two glaucoma specialists labeled the notes for each task, with a third glaucoma specialist serving as an adjudicator. We approached adjudication conservatively, flagging labels that were not exact matches for adjudication. We evaluated four medication-related extraction tasks – current topical medications (CTM), proposed changes to topical medications (ΔTM), current oral medications (COM), and proposed changes to oral medications (ΔOM). For topical medication tasks, labels were created for each eye, while labels were at the subject level for oral medication tasks. For CTM, labels consisted of name and frequency for each medication. For ΔTM, labels included these components as well as a change term (either “start”, “stop”, “increase”, or “decrease”) and duration if mentioned. For COM, labels consisted of medication name, dosage (if mentioned), and frequency. For ΔOM, labels also included a change term and duration if mentioned. Only prescription ophthalmic medications were included (i.e., over-the-counter medications such as artificial tears were excluded). A change from a preserved to preservative-free formulation of a medication was defined as a change, as such changes typically occur due to adverse effects typically associated with reduced medication adherence. However, a brand-generic switch was not considered a change, as such alterations are typically due to financial issues (e.g., insurance coverage).

Medications were not considered to be current if they had been discontinued for more than seven days. If there was no medication use, graders labelled as “None”. If medication use was unclear, the graders labelled as “Unspecified”. If laterality was unclear for CTM and ΔTM tasks, the grader was asked to add the phrase “(laterality unknown)” to the label of each eye. If multiple medications were present, details of each medication were comma-separated in the final label. This process established a ground truth benchmark for subsequent model performance assessment.

We split the dataset into development (10%), validation (10%), and held-out test sets (80%) stratified by clinician. Unlike traditional supervised machine learning workflows, the underlying LLM was not fine-tuned. Rather, the development set was used exclusively for prompt engineering and selection of few-shot examples rather than parameter optimization, requiring a relatively small dataset. We then used the validation set to refine prompt structure and rules before final evaluation. This process provided a large held-out test set that was not accessed during prompt development. We prioritized a large test set to provide a more stable estimate of performance. Prompts were frozen prior to final assessment with the test set. This approach is consistent with prompt-engineering frameworks in which model behavior is optimized through iterative prompt-refinement and evaluation rather than through parameter optimization.^18–20^

### Prompt Engineering and LLM Access

GLLaucoMed, a staged LLM pipeline for medication extraction, consisted of a structured agentic workflow with up to three sequential stages (initial extraction, validation, and conditional revision), followed by a standardization step.^14, 21, 22^ This pipeline incorporated rule-based instruction prompting, hierarchical evidence prioritization, few-shot examples, checklist-guided self-verification, and structured reasoning decomposition inspired by chain-of-thought prompting. Such approaches have been essential in minimizing hallucinations among LLM extraction pipelines.^23–25^ Initial input consisted of the clinical note and corresponding encounter date. The first stage generated initial output with supporting citations and reasoning. The second stage validated the extracted output against the original note utilizing the extracted citation and reasoning for additional context. Finally, the third stage was conditionally triggered only when validation did not pass. If validation passed, the initial output was retained, and the revision stage was skipped. If validation failed, the model was given the original note, the initial labels, and initial rationale, and was prompted to revise the output using the same prompt text provided in the first stage. Invalid outputs were coded as failed responses. Output components were equivalent to those requested of the graders. The full text of all prompts is available in the **Supplemental Material** (available at https://www.ophthalmologyscience.org).

To ensure consistent and machine-readable outputs, models were prompted to return responses in a structured JavaScript Object Notation (JSON) format. JSON extraction was performed first by direct parsing of the complete model response. In case of failure, we used a regular expression fallback to extract the final JSON object from the model response. Parsed outputs were stored along with task-level summaries, validation outputs, revision status, and reasoning fields. For CTM and ΔTM tasks, output included separate structured labels for right and left eyes (i.e., two labels per note). For COM and ΔOM tasks, output was at the subject level (i.e., one label per note).

We accessed LLMs within a secure cloud-based environment accessed using Microsoft Azure AI Foundry (Microsoft, Redmond, WA). All inference tasks were executed within HIPAA-compliant containers configured for PHI protection per a business associate agreement executed between Microsoft and the University of Miami. Amongst the available models in the foundry, we chose five leading LLMs: GPT-5.2, Claude Opus 4.6, DeepSeek-V3.2, Grok 4.1 fast non-reasoning, and Qwen3.6-35B-A3B. GPT-5.2 was routed through an Azure OpenAI client, Claude Opus 4.6 was routed through an Anthropic Foundry client, and DeepSeek, Grok, and Qwen were routed through OpenAI-compatible clients using model-specific endpoints. Model configuration included the endpoint, deployment name, model name, provider type, and supported reasoning-effort settings. We ran all model experiments with the same parameters, with temperature, top_p, and maximum token length set to 0.4, 0.9, and 750, respectively.

To equitably evaluate each model and avoid varied available reasoning levels amongst models, we set the reasoning effort of all models to “none”. Final outputs were written to model- and task-specific result files for subsequent evaluation.

### Label and Output Standardization

Given the free-text nature of labels and LLM outputs, we applied a text standardization step to enable subsequent text string comparison. We created an independent Python standardization script to standardize both the adjudicated labels and the LLM-generated output prior to performance evaluation. The script standardized the order of the output in the following order: change term (if applicable), medication name (generic), dosage (if applicable), frequency, and duration (if available). Any extraneous verbiage was eliminated. The script included a canonical library of medications and their brand names, which used this library to map medications to corresponding generic names. If a medication was not found in the canonical library, the script utilized an application programming interface (API) call to the Drugs@FDA database (https://www.accessdata.fda.gov/scripts/cder/daf/index.cfm) to obtain the generic name for a brand name medication. This step provided additional generalizability. Frequency terms were also standardized using a set canon (e.g., BID, TID, QID). Typographical errors were handled using a fuzzy matching algorithm employing the Damerau-Levenshtein distance, which measures the number of operations needed to transform one string into another.^26, 27^ Operations include insertions, deletions, substitutions, and transpositions of two adjacent characters. Medications were alphabetized and organized by change term (for ΔTM and ΔOM tasks). The script also allowed the structured parsing of the various output components (e.g., change term, name, frequency) for subsequent comparison. The standardization script can be accessed on GitHub: https://github.com/swarupswaminathan/llmnotes/tree/main/med_standardization.

### Statistical Analysis

We analyzed intergrader agreement for each task using Gwet AC1 given potential class imbalance. Labels were binarized for each task as “No” if the label consisted of “none” or “unspecified” and “Yes” if the label contained discrete medication data pertinent to that task (e.g., “latanoprost QHS” for CTM, “start timolol daily” for ΔTM). For positive cases, we also calculated inter-grader agreement with Jaccard Index (JI). JI measures the similarity between two sets as the ratio of their intersection to their union and is commonly used in data science to quantify the overlap between predicted and reference classifications.^28, 29^ Values range from 0 (no common elements) to 1 (identical elements). In this scenario, the token of comparison was the compound medication/frequency or medication/dosage/frequency phrase. For example, “latanoprost QHS” vs. “timolol daily” would have a JI of 0, while “travoprost QHS” vs. “dorzolamide/timolol BID, travoprost QHS” would have a JI of 0.5 (one of two medication/frequency entities present), and “methazolamide 25mg BID” and “methazolamide 25mg BID” would have a JI of 1. We utilized a compound token approach instead of a word-level token given the potential for false positives with the latter (e.g., “brimonidine TID, dorzolamide BID” vs. “brimonidine BID, dorzolamide TID” would falsely generate a perfect JI of 1 with word-level tokens). Of note, JI is a monotonic transformation of the F1 score.

To evaluate model performance, we binarized standardized, adjudicated labels and LLM outputs to define whether discrete medication data was present or absent pertinent to each task. We then used Gwet AC1 to quantify agreement between labels and LLM outputs. We calculated specificity, sensitivity (recall), precision, and F1 scores to characterize how well LLMs identified the presence of medication data. Among the positive cases, we then assessed for accuracy in text output using exact match accuracy (i.e., fraction of cases with an exact match of the full text between label and LLM output) as well as JI. Amongst these positive cases, we also computed exact match accuracy and JI for individual components: medication name, frequency, dosage (for COM and ΔOM), and change term (for ΔTM and ΔOM). For F1 score and JI, we computed 95% confidence intervals (CIs) using 1,000 bootstrap resamples, while we reported 95% CIs for specificity, sensitivity, and precision using Wilson CIs. We tabulated failure rates for each model and task; failures were not included in performance metric calculations.

For CTM and ΔTM, we also converted labels and LLM outputs into number of topical medications or number of changes in topical medications. We then computed the mean absolute error (MAE) using the estimated number of current medications or change in medications from the LLM outputs compared to labels. To account for potential bias induced by unmedicated subjects, we computed these MAE values both overall and among positive cases only.

## Results

A total of 1,984 labels for CTM and ΔTM tasks and 992 labels for COM and ΔOM tasks were evaluated in the test set. Gwet AC1 for intergrader agreement was 0.799, 0.888, 0.985, and 0.988 for CTM, ΔTM, COM, and ΔOM, respectively. Among positive cases, inter-grader JI was 0.811, 0.622, 0.745, and 0.621, respectively. A total of 1,354 eyes (68.2%) were using topical medications per graded labels, while 404 eyes (20.3%) had changes noted regarding their topical medication regimens. The mean number of current topical medications was 1.3±1.2 (range 0-7) and mean number of changes in topical medications was 0.3±0.7 (range 0-7; **Figure 1**). Only 47 subjects (4.7%) were on oral medications, while 25 subjects (2.5%) had changes noted regarding oral medications.

**Figure 1.**
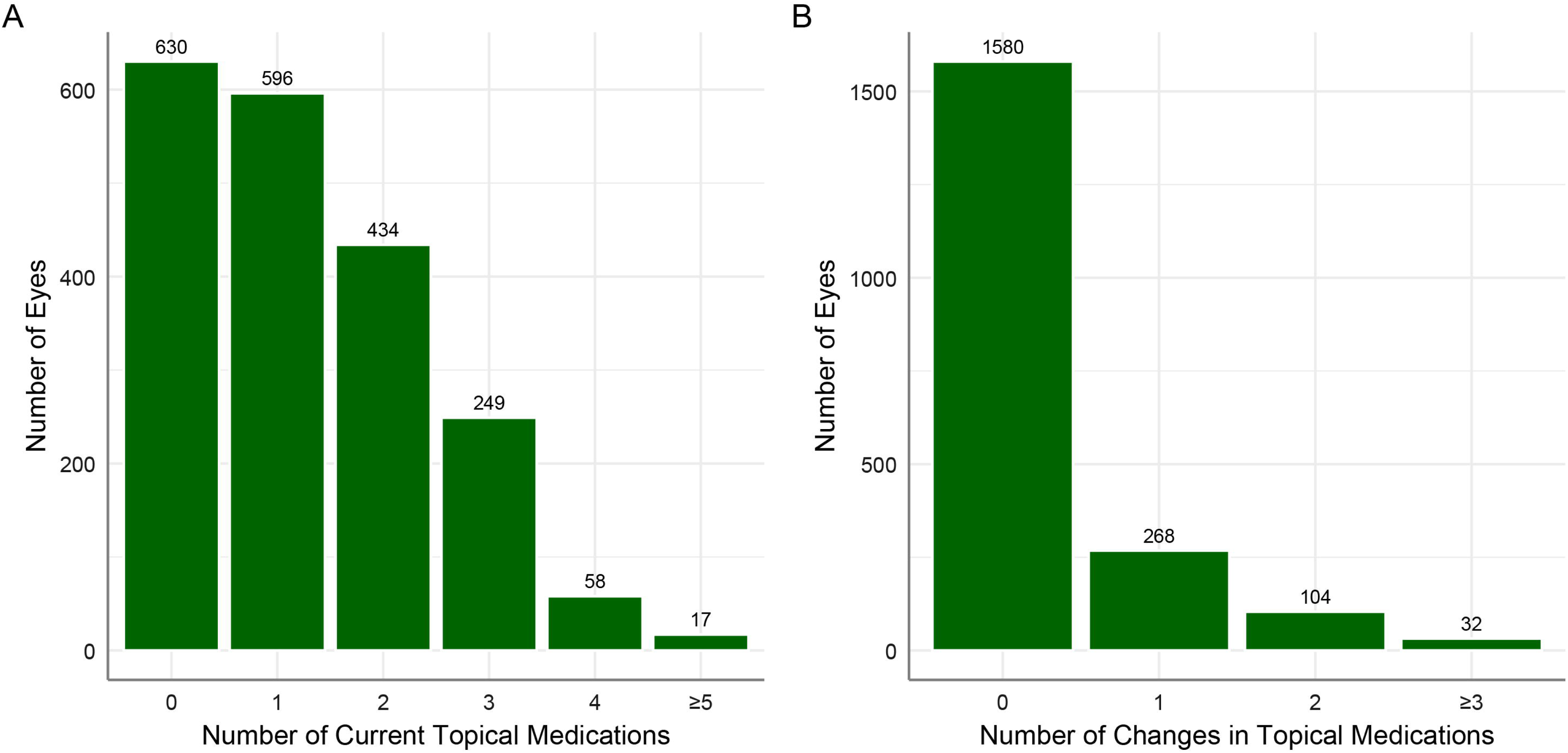
Histogram demonstrating distribution of number of (A) current topical medications and (B) changes in topical medications.

Model performance in identifying the presence or absence of medication data pertinent to each task in notes is shown in **Table 1**. Failures were extremely rare, ranging from 0 notes (0%) to 6 notes (0.006%) among the various models and tasks. For CTM, Gwet AC1 ranged from 0.772 (Qwen) to 0.898 (Claude). Claude had the highest F1 score at 0.985 (95% CI: 0.980-0.989). MAE for the number of current topical medications ranged from 0.061 (Claude) to 0.153 (Qwen). Distribution of errors is displayed in **Figure 2A**. Sensitivity of detecting the use of topical medications ranged from 0.957 (Grok) to 0.990 (Claude). Precision ranged from 0.970 (GPT) to 0.980 (Claude). For ΔTM, Gwet AC1 ranged from 0.889 (Grok) to 0.931 (GPT). Claude had the highest F1 score at 0.905 (95% CI: 0.883-0.925). MAE for the number of changes in topical medications ranged from 0.074 (GPT) to 0.130 (Grok). Distribution of errors is displayed in **Figure 2B**. Sensitivity of detecting a change in topical medications ranged from 0.775 (DeepSeek) to 0.941 (Claude). Precision ranged from 0.809 (Grok) to 0.894 (Qwen). For COM, Gwet AC1 ranged from 0.985 (Qwen) to 0.992 (Grok), and Grok had the highest F1 score at 0.923 (95% CI: 0.860-0.972). For ΔOM, Gwet AC1 ranged from 0.989 (Grok) to 0.997 (Claude), with Claude having the highest F1 score at 0.958 (95% CI: 0.889-1.000).

**Table 1.** Model performance metrics for binary classification utilizing different large language models for medication-related tasks

|  |  | Claude | DeepSeek | GPT | Grok | Qwen |
| --- | --- | --- | --- | --- | --- | --- |
| **CTM** | Failure rate | 0% | 0% | 0.001% (S-1) | 0% | 0% |
|  | Gwet AC1 | 0.898 (0.885 – 0.912) | 0.822 (0.805 - 0.840) | 0.858 (0.842 - 0.873) | 0.840 (0.823 - 0.855) | 0.772 (0.753 - 0.791) |
|  | Specificity | 0.956 (0.937 – 0.969) | 0.948 (0.927 – 0.962) | 0.935 (0.913 – 0.952) | 0.956 (0.937 – 0.969) | 0.938 (0.916 – 0.954) |
|  | Sensitivity (recall) | 0.990 (0.983 – 0.994) | 0.966 (0.955 – 0.974) | 0.987 (0.979 – 0.992) | 0.957 (0.945 – 0.967) | 0.968 (0.957 – 0.976) |
|  | Precision | 0.980 (0.971 - 0.986) | 0.975 (0.966 - 0.982) | 0.970 (0.960 - 0.978) | 0.979 (0.970 - 0.985) | 0.971 (0.961 - 0.979) |
|  | F1 score | 0.985 (0.980 - 0.989) | 0.971 (0.964 - 0.977) | 0.978 (0.973 - 0.984) | 0.968 (0.961 - 0.975) | 0.970 (0.963 - 0.976) |
|  | MAE | 0.061 (0.048 - 0.076) | 0.123 (0.105 - 0.141) | 0.088 (0.072 - 0.104) | 0.136 (0.117 - 0.158) | 0.153 (0.133 - 0.172) |
| **ΔTM** | Failure rate | 0.003% (S-3) | 0.003% (S-3) | 0.003% (S-5) | 0.009% (S-6, M-3) | 0.006% (S-6) |
|  | Gwet AC1 | 0.926 (0.915 - 0.937) | 0.901 (0.887 - 0.914) | 0.931 (0.921 - 0.942) | 0.889 (0.875 - 0.903) | 0.909 (0.896 - 0.922) |
|  | Specificity | 0.964 (0.953 - 0.972) | 0.973 (0.964 – 0.980) | 0.980 (0.972 – 0.986) | 0.947 (0.934 – 0.957) | 0.974 (0.965 – 0.981) |
|  | Sensitivity (recall) | 0.941 (0.913 – 0.960) | 0.775 (0.732 – 0.813) | 0.874 (0.838 – 0.903) | 0.873 (0.837 – 0.902) | 0.834 (0.795 – 0.867) |
|  | Precision | 0.872 (0.837 - 0.900) | 0.884 (0.847 - 0.913) | 0.917 (0.885 - 0.941) | 0.809 (0.769 – 0.843) | 0.894 (0.859 - 0.921) |
|  | F1 score | 0.905 (0.883 - 0.925) | 0.826 (0.797 - 0.855) | 0.897 (0.875 - 0.919) | 0.842 (0.815 – 0.868) | 0.865 (0.837 - 0.889) |
|  | MAE | 0.077 (0.062 - 0.091) | 0.123 (0.103 - 0.143) | 0.074 (0.059 - 0.089) | 0.130 (0.110 - 0.152) | 0.093 (0.077 - 0.109) |
| **COM** | Failure rate | 0% | 0% | 0% | 0% | 0% |
|  | Gwet AC1 | 0.991 (0.985 - 0.996) | 0.986 (0.978 - 0.993) | 0.987 (0.979 - 0.994) | 0.992 (0.986 - 0.997) | 0.985 (0.977 - 0.992) |
|  | Specificity | 0.997 (0.991 – 0.999) | 0.997 (0.991 – 0.999) | 0.997 (0.991 – 0.999) | 0.999 (0.994 – 1.000) | 0.994 (0.986 – 0.997) |
|  | Sensitivity (recall) | 0.915 (0.801 – 0.966) | 0.851 (0.723 – 0.926) | 0.872 (0.748 – 0.940) | 0.851 (0.723 – 0.926) | 0.915 (0.801 – 0.966) |
|  | Precision | 0.935 (0.825 – 0.978) | 0.930 (0.814 – 0.976) | 0.932 (0.818 – 0.977) | 0.976 (0.874 – 0.996) | 0.878 (0.758 – 0.943) |
|  | F1 score | 0.923 (0.860 – 0.972) | 0.887 (0.812 – 0.947) | 0.899 (0.825 – 0.956) | 0.906 (0.837 – 0.966) | 0.894 (0.824 – 0.954) |
| **ΔOM** | Failure rate | 0% | 0.004% (M-4) | 0% | 0.001% (M-1) | 0% |
|  | Gwet AC1 | 0.997 (0.993 - 1.000) | 0.991 (0.985 - 0.996) | 0.996 (0.992 - 0.999) | 0.989 (0.982 - 0.995) | 0.994 (0.988 - 0.998) |
|  | Specificity | 0.999 (0.994 – 1.000) | 0.999 (0.994 – 1.000) | 0.999 (0.994 – 1.000) | 0.999 (0.994 – 1.000) | 0.998 (0.992 – 0.999) |

|  | Claude | DeepSeek | GPT | Grok | Qwen |
| --- | --- | --- | --- | --- | --- |
| Sensitivity (recall) | 0.960 (0.805 – 0.993) | 0.720 (0.524 – 0.857) | 0.920 (0.750 – 0.978) | 0.750 (0.551 – 0.880) | 0.960 (0.805 – 0.993) |
| Precision | 0.960 (0.805 – 0.993) | 0.947 (0.754 - 0.991) | 0.958 (0.798 - 0.993) | 0.947 (0.754 – 0.991) | 0.923 (0.759 – 0.979) |
| F1 score | 0.958 (0.889 – 1.000) | 0.815 (0.681 - 0.923) | 0.937 (0.851 - 1.000) | 0.835 (0.694 – 0.943) | 0.940 (0.857 – 1.000) |

CTM = current topical medications; ΔTM = proposed changes to topical medications; COM = current oral medications; ΔOM = proposed changes to oral medications; MAE = mean absolute error; MSE = mean squared error. Failure-rate notation: S = standardization failure; M = model failure, with the number indicating the number of notes that failed for each etiology.

**Figure 2.**
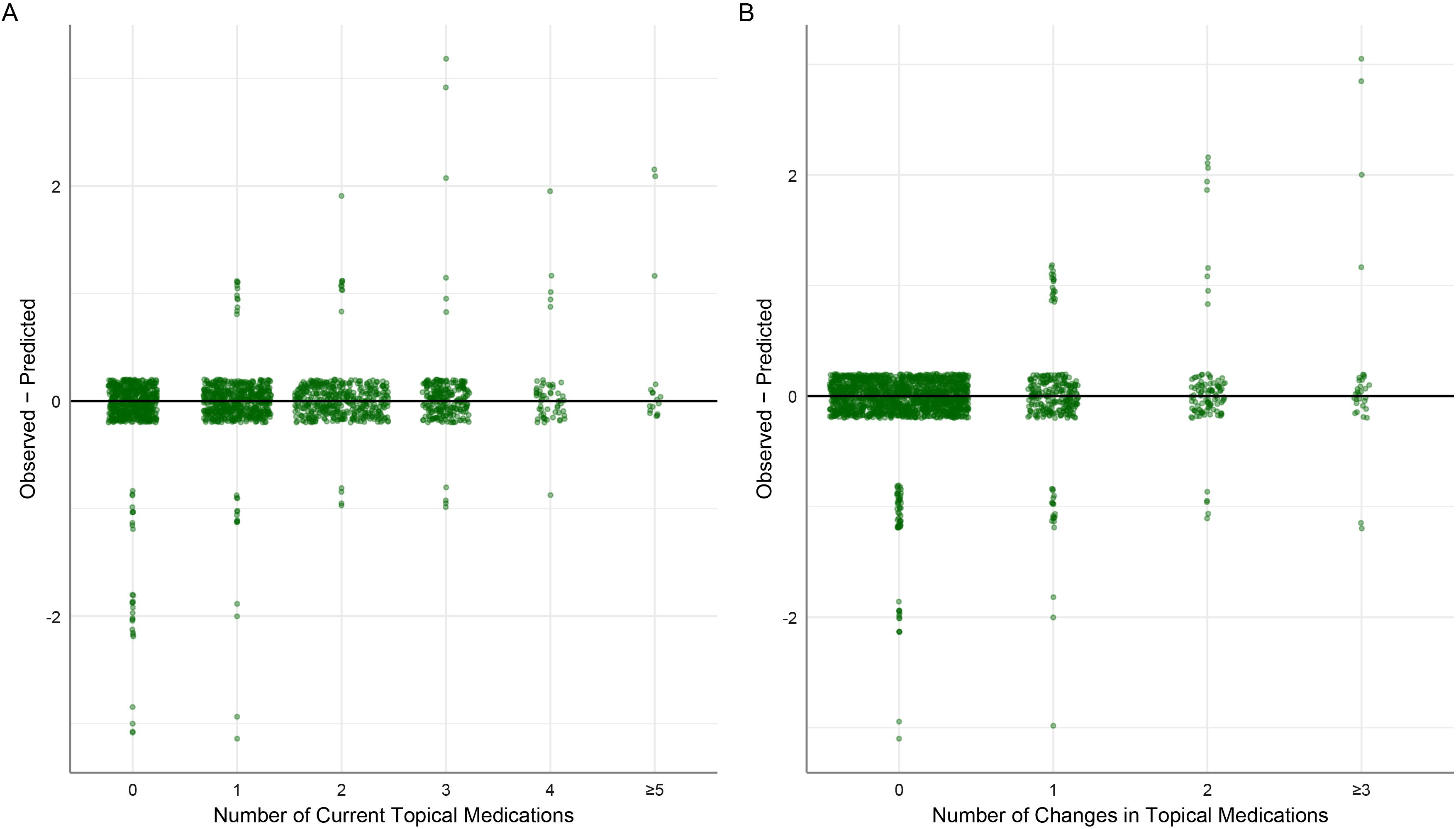
Sina plots demonstrating the relationship between (A) the number of current topical medications and the difference between ground truth label and predicted value, and (B) the number of changes in topical medications and the difference between ground truth label and predicted value. Predictions from Claude were utilized. Horizontal point spread is proportional to local density. Points are jittered for visibility.

Model performance for text analysis of positive cases only is shown in **Table 2**. For CTM, Claude had the highest exact match accuracy at 0.882 (95% CI: 0.864-0.898) and highest JI at 0.918 (95% CI: 0.905-0.931). Positive-case MAE for number of topical medications ranged from 0.053 (Claude) to 0.178 (Qwen). For ΔTM, Claude had the highest exact match accuracy at 0.780 (95% CI: 0.737-0.817) and highest JI at 0.827 (95% CI: 0.796-0.860). Positive-case MAE for change in number of topical medications ranged from 0.197 (Claude) to 0.476 (DeepSeek). For COM, Claude had the highest exact match accuracy at 0.8723 (95% CI: 0.7483-0.9402), while Grok had the highest JI at 0.8516 (95% CI: 0.7447-0.9362). For ΔOM positive cases, Claude had the highest JI at 0.922 (95% CI: 0.800-1.000) and the highest exact match accuracy at 0.920 (95% CI: 0.750-0.978).

**Table 2.** Model performance metrics of text analysis among positive cases.

|  |  | **Claude** | **DeepSeek** | **GPT** | **Grok** | **Qwen** |
| --- | --- | --- | --- | --- | --- | --- |
| **CTM** | Jaccard Index | 0.918 (0.905 - 0.931) | 0.848 (0.830 - 0.863) | 0.896 (0.881 - 0.910) | 0.858 (0.841 - 0.874) | 0.809 (0.789 - 0.828) |
| **N=1354** | Exact match accuracy | 0.882 (0.864 - 0.898) | 0.791 (0.769 - 0.812) | 0.861 (0.842 - 0.879) | 0.795 (0.772 - 0.815) | 0.730 (0.705 - 0.753) |
|  | MAE | 0.053 (0.039 - 0.068) | 0.137 (0.115 - 0.160) | 0.082 (0.064 - 0.103) | 0.166 (0.140 - 0.191) | 0.178 (0.153 - 0.203) |
| **ΔTM** | Jaccard Index | 0.827 (0.796 - 0.860) | 0.668 (0.626 - 0.711) | 0.781 (0.743 - 0.818) | 0.722 (0.681 - 0.765) | 0.714 (0.672 - 0.756) |
| **N=404** | Exact match accuracy | 0.780 (0.737 - 0.817) | 0.619 (0.571 - 0.665) | 0.749 (0.704 - 0.789) | 0.670 (0.622 - 0.714) | 0.659 (0.612 - 0.704) |
|  | MAE | 0.197 (0.151 - 0.250) | 0.476 (0.396 - 0.559) | 0.275 (0.210 - 0.344) | 0.345 (0.271 - 0.420) | 0.325 (0.267 - 0.389) |
| **COM** | Jaccard Index | 0.870 (0.766 – 0.957) | 0.765 (0.638 - 0.872) | 0.788 (0.660 - 0.894) | 0.852 (0.745 - 0.936) | 0.809 (0.702 – 0.915) |
| **N=47** | Exact match accuracy | 0.872 (0.748 - 0.940) | 0.766 (0.628 - 0.864) | 0.787 (0.651 - 0.880) | 0.851 (0.723 - 0.926) | 0.808 (0.675 - 0.896) |
| **ΔOM** | Jaccard Index | 0.922 (0.800 - 1.000) | 0.677 (0.480 - 0.840) | 0.877 (0.760 - 1.000) | 0.583 (0.375 - 0.792) | 0.841 (0.680 – 0.960) |
| **N=25** | Exact match accuracy | 0.920 (0.750 - 0.978) | 0.680 (0.484 - 0.828) | 0.880 (0.700 - 0.958) | 0.583 (0.388 - 0.755) | 0.840 (0.653 – 0.936) |

Positive cases were defined as reference-standard labels that were not None or Unspecified.
CTM = current topical medications; ΔTM = proposed changes to topical medications; COM = current oral medications; ΔOM = proposed changes to oral medications; MAE = mean absolute error.

Model performance for individual components of the labels of positive cases is shown in **Table 3**. When evaluating medication names, Claude had the highest exact match accuracy at 0.948 (95% CI: 0.934-0.958) and highest JI at 0.968 (95% CI: 0.960-0.976) for CTM. For ΔTM, Claude had the highest exact match accuracy at 0.851 (95% CI: 0.814-0.883) and highest JI at 0.891 (95% CI: 0.862-0.917). For COM, Claude and Qwen both had the highest exact match accuracy of 0.915 (95% CI: 0.801-0.966), while Qwen had the highest JI at 0.917 (95% CI: 0.830-0.979). For ΔOM, Claude and Qwen both had the highest exact match accuracies of 0.920 (95% CI: 0.750-0.978), while Qwen had the highest JI at 0.941 (95% CI: 0.840-1.000).

**Table 3.** Model performance metrics when evaluating individual components of positive cases.

|  | Claude | DeepSeek | GPT | Grok | Qwen |
| --- | --- | --- | --- | --- | --- |
| **CTM** |  |  |  |  |  |
| ***Medication name*** |  |  |  |  |  |
| Jaccard Index | 0.968 (0.960 - 0.976) | 0.914 (0.903 - 0.928) | 0.956 (0.947 - 0.965) | 0.909 (0.896 - 0.922) | 0.891 (0.877 - 0.905) |
| Exact match accuracy | 0.948 (0.934 - 0.958) | 0.869 (0.850 - 0.886) | 0.928 (0.913 - 0.940) | 0.856 (0.836 - 0.874) | 0.825 (0.804 - 0.844) |
| ***Frequency*** |  |  |  |  |  |
| Jaccard Index | 0.943 (0.932 - 0.954) | 0.888 (0.872 - 0.902) | 0.931 (0.920 - 0.943) | 0.893 (0.879 - 0.906) | 0.866 (0.850 - 0.881) |
| Exact match accuracy | 0.823 (0.802 - 0.843) | 0.755 (0.731 - 0.777) | 0.801 (0.779 - 0.822) | 0.748 (0.724 - 0.771) | 0.712 (0.687 - 0.735) |
| **ΔTM** |  |  |  |  |  |
| ***Medication name*** |  |  |  |  |  |
| Jaccard Index | 0.891 (0.862 - 0.917) | 0.711 (0.670 - 0.748) | 0.835 (0.801 - 0.867) | 0.807 (0.770 - 0.841) | 0.786 (0.746 - 0.823) |
| Exact match accuracy | 0.851 (0.814 - 0.883) | 0.663 (0.616 - 0.708) | 0.808 (0.767 - 0.844) | 0.757 (0.713 - 0.797) | 0.739 (0.694 - 0.779) |
| ***Frequency*** |  |  |  |  |  |
| Jaccard Index | 0.862 (0.831 - 0.892) | 0.700 (0.656 - 0.742) | 0.820 (0.785 - 0.856) | 0.768 (0.727 - 0.806) | 0.761 (0.724 - 0.800) |
| Exact match accuracy | 0.723 (0.677 - 0.764) | 0.589 (0.541 - 0.636) | 0.682 (0.635 - 0.725) | 0.645 (0.597 - 0.690) | 0.632 (0.584 - 0.678) |
| ***Change Term*** |  |  |  |  |  |
| Jaccard Index | 0.899 (0.871 - 0.925) | 0.723 (0.682 - 0.764) | 0.834 (0.799 - 0.865) | 0.804 (0.767 - 0.840) | 0.788 (0.750 - 0.826) |
| Exact match accuracy | 0.752 (0.708 - 0.792) | 0.594 (0.546 - 0.641) | 0.697 (0.650 - 0.739) | 0.675 (0.628 - 0.719) | 0.654 (0.606 - 0.699) |
| **COM** |  |  |  |  |  |
| **Medication name** |  |  |  |  |  |
| Jaccard Index | 0.914 (0.830 – 0.979) | 0.850 (0.745 – 0.936) | 0.872 (0.766 – 0.957) | 0.852 (0.745 – 0.936) | 0.917 (0.830 – 0.979) |
| Exact match accuracy | 0.915 (0.801 – 0.966) | 0.851 (0.723 – 0.926) | 0.872 (0.748 – 0.940) | 0.851 (0.723 – 0.926) | 0.915 (0.801 – 0.966) |
| **Frequency** |  |  |  |  |  |
| Jaccard Index | 0.871 (0.766 – 0.957) | 0.766 (0.638 – 0.872) | 0.809 (0.681 – 0.915) | 0.854 (0.745 – 0.957) | 0.828 (0.723 – 0.915) |
| Exact match accuracy | 0.872 (0.748 – 0.940) | 0.766 (0.628 – 0.864) | 0.809 (0.675 – 0.896) | 0.851 (0.723 – 0.926) | 0.830 (0.699 – 0.911) |
| **Dosage** |  |  |  |  |  |
| Jaccard Index | 0.913 (0.830 - 0.979) | 0.850 (0.745 - 0.957) | 0.850 (0.745 - 0.936) | 0.851 (0.745 - 0.936) | 0.874 (0.766 - 0.957) |
| Exact match accuracy | 0.915 (0.801 - 0.966) | 0.851 (0.723 - 0.926) | 0.851 (0.723 - 0.926) | 0.851 (0.723 - 0.926) | 0.872 (0.748 - 0.940) |
| **ΔOM** |  |  |  |  |  |
| **Medication name** |  |  |  |  |  |
| Jaccard Index | 0.940 (0.840 - 1.000) | 0.696 (0.500 - 0.860) | 0.900 (0.780 - 1.000) | 0.726 (0.542 - 0.896) | 0.941 (0.840 - 1.000) |
| Exact match accuracy | 0.920 (0.750 - 0.978) | 0.680 (0.484 - 0.828) | 0.880 (0.700 - 0.958) | 0.708 (0.508 - 0.851) | 0.920 (0.750 - 0.978) |
| **Frequency** |  |  |  |  |  |
| Jaccard Index | 0.938 (0.840 - 1.000) | 0.698 (0.520 - 0.860) | 0.899 (0.780 - 1.000) | 0.684 (0.500 - 0.854) | 0.901 (0.780 - 1.000) |
| Exact match accuracy | 0.920 (0.750 - 0.978) | 0.680 (0.484 - 0.828) | 0.880 (0.700 - 0.958) | 0.667 (0.467 - 0.820) | 0.880 (0.700 - 0.958) |
| **Dosage** |  |  |  |  |  |
| Jaccard Index | 0.938 (0.840 - 1.000) | 0.698 (0.520 - 0.860) | 0.901 (0.780 - 1.000) | 0.640 (0.458 - 0.812) | 0.901 (0.780 - 1.000) |
| Exact match accuracy | 0.920 (0.750 - 0.978) | 0.680 (0.484 - 0.828) | 0.880 (0.700 - 0.958) | 0.625 (0.427 - 0.788) | 0.880 (0.700 - 0.958) |
| ***Change Term*** |  |  |  |  |  |
| Jaccard Index | 0.940 (0.840 - 1.000) | 0.701 (0.520 - 0.880) | 0.899 (0.760 - 1.000) | 0.731 (0.542 - 0.875) | 0.941 (0.840 - 1.000) |
| Exact match accuracy | 0.920 (0.750 - 0.978) | 0.680 (0.484 - 0.828) | 0.880 (0.700 - 0.958) | 0.708 (0.508 - 0.851) | 0.920 (0.750 - 0.978) |

CTM = current topical medications; ΔTM = proposed changes to topical medications; COM = current oral medications; ΔOM = proposed changes to oral medications.

Regarding model performance for frequency, Claude performed best across all tasks, with exact match accuracy ranging from 0.723 to 0.920. For model performance of COM dosage, Claude had the highest exact match accuracy at 0.915 (95% CI: 0.801-0.966) and highest JI at 0.913 (95% CI: 0.830-979). For ΔOM dosage, Claude had the highest exact match accuracy at 0.920 (95% CI: 0.750-0.978) and highest JI at 0.938 (95% CI: 0.840-1.000). For model performance of ΔTM change term (e.g., “start”, “increase”), Claude had the highest exact match accuracy at 0.752 (95% CI: 0.708-0.792) and highest JI at 0.899 (95% CI: 0.871-0.925). For ΔOM change term, Claude and Qwen both had the highest exact match accuracies of 0.920 (95% CI: 0.750-0.978) and Qwen had the highest JI of 0.941 (95% CI: 0.840-1.000).

When reviewing a random selection of 50 failed matches among the various tasks, the main cause (50-80% across tasks) was ambiguity in the clinical notes regarding current or proposed regimens (e.g., contradictory information in two sections of the note, ambiguity regarding whether patient was using medications as prescribed, or ambiguity regarding whether medications were currently used or a proposed change), leading the labelers to use one data point for their labels while the LLM utilized the another data point for its output. True logical errors in LLM reasoning appeared to comprise only 12-20% of failures. **Supplementary Table 4** contains examples of failures (available at https://www.ophthalmologyscience.org).

## Discussion

In this study, the GLLaucoMed agentic workflow demonstrated strong performance for extracting medication information in a standardized fashion from free-text EHR glaucoma clinical notes in a secure environment. Approximately 6 out of 10 eyes were currently on topical medications, while clinicians proposed changes in topical medication regimens for approximately 2 out of 10 eyes in this cross-sectional analysis. Across the four tasks, multiple LLMs achieved high exact match accuracy and JI values when compared with expert-labeled reference standards. Overall, Claude appeared to be the strongest LLMs across all tasks. The low MAE in estimates of the number of topical medications or change in topical medications reflects the high accuracy of this pipeline if outputs were quantified, which may be valuable to research applications. Use of the Microsoft Azure AI Foundry provided access to leading LLMs within secure containers to ensure no compromise of potential PHI. By using an agentic workflow followed by output standardization, the GLLaucoMed pipeline can provide rapid and accurate assessments of unstructured clinical notes to fully characterize the treatment intensity of glaucoma patients within EHR-derived datasets.

Model performance varied by task and label component. Claude and GPT were consistently among the strongest-performing models across extraction tasks, consistent with their relative standing on established third-party evaluation platforms, in which both models rank among the highest-performing systems across a broad range of language understanding benchmarks.^30^ However, these results do not provide model-internal explanations for task-specific performance differences, and therefore, we did not entertain causal conclusions regarding why a particular model performed better on a specific task.

The ability of LLMs to identify changes in topical treatment is a key aspect of this work. When a treatment change occurs, can the LLM identify the change and fully characterize it? Sensitivity for ΔTM (i.e., how well the model correctly identified that a treatment change occurred) was 0.941 for Claude and 0.874 for GPT, while precision (i.e., how often a change had truly occurred when the model predicted a change) was 0.872 for Claude and 0.917 for GPT. When evaluating the text of positive cases, exact match accuracy and JI were 0.780 and 0.827 for Claude and 0.749 and 0.781 for GPT, respectively. These values were lower compared to those of the other three tasks, potentially due to the complex nature of distinguishing change within a clinical note, which can be challenging given potential contradictory information in different sections of the note or ambiguity regarding current medications (**Supplemental Table 4**).

Analysis of multiple chronological notes per subject may improve model performance. Performance was also more variable across models for the CTM and ΔTM tasks. Again, this difference in performance may be attributable to the greater diversity and number of topical medications compared to oral medications, resulting in greater complexity.

Among positive cases, medication name extraction consistently outperformed frequency extraction, particularly for topical medication tasks. For CTM, Claude achieved exact match accuracies of 0.948 and 0.823 for medication name and frequency, respectively, while for ΔTM, Claude achieved exact match accuracies of 0.851 and 0.723 for medication name and frequency, respectively. This distinction carries meaningful clinical implications – while medication name recognition alone is insufficient for complete treatment characterization, accurate identification of a medication name or a change in medication name is more likely to be considered a key detail by clinicians compared to capturing frequency or a change in frequency.

Frequency data is of limited interpretive value without the corresponding medication name, whereas a correctly identified medication name retains some clinical utility even in the absence of the associated frequency. The comparatively lower performance on frequency extraction may reflect the greater variability in how frequency was documented in clinical text; it may be omitted from treatment plans or contradictory information may be present within the note.

Prior NLP approaches in ophthalmology applied supervised named entity recognition (NER) methods to glaucoma medication extraction from clinical notes.^12^ Majid et al. demonstrated strong performance of LLMs for ophthalmic medication NER by extracting medication name, route, laterality, and frequency.^13^ However, the unit of analysis in this study was an isolated line of text rather than the entire clinical note. A subsequent study benchmarking GPT-5, LLaMA 3.1–70B, and Mistral 7B expanded LLM-based ophthalmology extraction to full progress notes and multiple clinical entity categories, but its target entities were broad ophthalmic findings rather than medication regimens.^31^ We previously reported analyses of risk factors for glaucoma progression within clinical notes but using a locally installed LLM, which could serve as an alternate albeit less powerful solution.^17^ The present study differs from these prior works by providing full clinical notes to leading LLMs in a secure cloud environment and evaluating whether they can extract not only current medications but also proposed medication changes from full notes – a clinically meaningful advance beyond prior extraction studies. A complete note is the more appropriate unit of analysis for clinical application rather than an isolated line of text. This design allowed the GLLaucoMed pipeline to capture clinically actionable treatment-plan information in a comprehensive fashion. Use of a staged extraction, validation, and revision pipeline optimized accuracy. If extracted locally by contributing members, such standardized data derived from unstructured notes could serve as additional visit-level data submitted to large data repositories such as the AAO IRIS Registry. Access to such data would substantially strengthen the quality of research initiatives possible with these databases.

The standardization step in our pipeline was another strength of this study. Model outputs and expert labels were not compared in raw free-text form but rather in a standardized format. The script mapped brand to generic names, normalized frequencies and durations, removed extra verbiage, and used well-established fuzzy matching methods to address typographical errors. The script featured key flexibility in also being able to access the FDA drug database. Standardization was essential given significant variation in how medication names were written and frequencies were expressed. The low number of failures supports the feasibility of pairing LLM extraction with deterministic post-processing before quantitative evaluation.

Several limitations should be acknowledged. First, the study was conducted using clinical notes from a single institution, which may limit generalizability to other ophthalmology practices that feature different documentation styles. However, it is worth noting that notes were drawn from twenty different BPEI glaucoma specialists, representing diversity in note styles. Only one section of the prompts (describing a unique frequency notation convention used at BPEI) and the reference section (containing acronyms and abbreviations) were institution-specific. Second, successful deployment of these LLMs requires access to the Azure AI Foundry. While access may be limited currently to larger academic institutions, we believe that this barrier may be overcome as accessibility increases over time. Finally, although the standardization script reduced variation in medication naming and formatting, the final performance metrics depended on the accuracy of both model extraction and post-processing. Future efforts may focus on validating the GLLaucoMed workflow at other institutions.

Prompts can be modified to include institution-specific patterns, particularly with a “few-shot” approach to capture institution-specific writing styles. The prompts in this work may serve as the foundation for subsequent prompt refinement. This approach can also be extended to other areas in ophthalmology to capture key data from unstructured notes. Longitudinal assessments of medication status using multiple notes per subject may also be considered.

In summary, the GLLaucoMed pipeline demonstrated strong performance for extracting structured glaucoma medication information from unstructured EHR clinical notes in a secure environment. This agentic workflow performed particularly well for medication name extraction and quantification of medications. These findings support the potential role of secure LLM agentic workflows for transforming free-text clinical notes into structured medication data for further research use.

**Financial Support:** NIH EY036593 (FAM), NIH K23 EY033831 (SSS)

## Data Availability

Data are available upon reasonable request, excluding notes given the potential inclusion of protected health information.

https://github.com/swarupswaminathan/llmnotes/tree/main/med_standardization

## Abbreviations

API: Application Programming Interface
EHR: Electronic Health Record
LLM: Large Language Model
NLP: Natural language processing
PHI: Protected Health Information.

