## Supplemental Table 4 for "GLLaucoMed: A Secure LLM-Powered Agentic Workflow for Automated Medication Extraction from Free-Text Glaucoma Clinical Notes"

**Supplemental Table 4.** Examples of inaccurate LLM responses. Responses are for one eye for topical medication tasks. Pertinent note text is highlighted in red.

| Task | Note Text | Adjudicated label | LLM output | Claude LLM reasoning |
|---|---|---|---|---|
| CTM | Impression: 1. Primary open angle glaucoma OD IOP improved after increasing azopt to BID (also on xal 1/1) - patient requesting changing azopt to trusopt due to insurance issues Per paper records, Tmax 28/45 CCT 511/575 Family history: brother and aunt with glaucoma (aunt blind) HVF OD (**/**/****): nonspecific inferior depression (significant movement on eyetracker) OCT RNFL (**/**/****): overall thickness 70 microns, borderline superior thinning (stable compared to **/****) 2. PBK with K scar 2/2 bacterial keratitis (MRSA) Note in paper chart from **/**/**** in resident cornea clinic with note stating poor prognosis for PK OS (Dr. ***) Patient is not interested in further surgical intervention, denies pain or vision changes Bscan today (**/**/****): No RD or mass, significant cupping OU Notes occasional irritation but no pain, no pain today 3. Pseudophakia OU Plan 1. Continue trusopt 2/2, and xal 1/1 (refilled for 1 year today) 2. Artificial tears for irritation 3. Monocular precautions 4. Return in 3-4 months IOP and repeat HVF OD | brinzolamide BID, latanoprost QHS | dorzolamide BID, latanoprost QHS | {"task1_reasoning": "Priority 2: The Plan section contains the trigger word 'continue' for trusopt 2/2 and xal 1/1. The Impression section mentions the patient was on azopt and xal, and is requesting a change to trusopt. The Plan confirms the switch has been made by stating 'Continue trusopt 2/2, and xal 1/1'. Trusopt is expanded to dorzolamide; xal is expanded to latanoprost. Per the prostaglandin QHS rule, latanoprost 1/1 becomes QHS for each eye. Trusopt 2/2 = dorzolamide BID each eye. Artificial tears are excluded as OTC.", "task1_citation": "OD Citation: Continue trusopt 2/2, and xal 1/1 OS Citation: Continue trusopt 2/2, and xal 1/1", "task2_reasoning": "OD Reason: OS Reason: ", "task3_reasoning": null, "task3_citation": null} |

| | | | | |
|---|---|---|---|---|
| CTM | Summary: Referred by Dr. *** for hypotony OD. - Outside glaucoma specialist: MD on Wisconson - no longer seeing - Surg hx: Trab OD; CEIOL OU - Laser hx: SLT OD - Glaucoma FHx: none - Tmax: high 20s OU - CCT / - Gonio: open OU (11/2024) - Target IOP: mid teens OU - Med AEs: n/a **/**/****: IOP 3 / 17 On Xal 0/1; Tim 0/1; PF 4/0 -- at target OU Imaging: HVF 24-2 (**/****) -- high FP, SNS/INS - base OD reliable, inf defect - base OS OCT RNFL (**/****) -- inf thinning - base OD sup/inf thinning - base OS OCT GCIPL (**/****) -- diffuse thinning - base OD diffuse thinning - base OS ONH photo pending Assessment: 1. Primary open angle glaucoma moderate OU IOP acceptable OS. 2. Hypotony OD No bleb leak on exam. IOP still low OD. IOP not improved with Durezol. Discussed trab revision OD to increase IOP. RBA discussed with patient and patient agrees to probeed. 3. Dry Eye Syndrome OD Tearing OD due to dryness and bleb. Plan: 1. Continue Timolol OS QAM and Latanoprost OS QHS 2. Continue Durezol OD QID 3. PFATs OD QID and gel drops OD QHS Return for trab revision OD | prednisolone QID | difluprednate QID | {"task1_reasoning": "Priority 1: The current encounter section dated **/**/**** (within day of **/**/****) lists medications: 'Xal 0/1; Tim 0/1; PF 4/0'. Xal 0/1 = Xalatan (latanoprost) 0 OD / 1 OS latanoprost QHS OS (prostaglandin rule). Tim 0/1 = timolol 0 OD / 1 OS timolol daily OS. PF 4/0 = Pred Forte 4 OD / 0 OS Pred Forte QID OD. However, the Plan section says 'Continue Durezol OD QID', which indicates Durezol is also a current medication OD. Looking back at the note, the statement 'IOP not improved with Durezol' confirms the patient is currently using Durezol OD. The Plan 'continue' trigger word confirms Durezol OD QID is current. PF 4/0 likely refers to Pred Forte QID OD from the encounter notation. However, re-examining: the patient has hypotony OD and is using Durezol OD to try to raise IOP. The 'PF 4/0' notation could mean Pred Forte 4x/day OD. But the Plan says 'Continue Durezol OD QID' - this suggests Durezol is the steroid being used. It's possible PF 4/0 actually refers to Pred Forte and Durezol is a separate medication. Since both appear to be current based on the encounter data and Plan 'continue' trigger, I include both. Actually, reconsidering: PF 4/0 in the context where the Plan says continue Durezol QID OD - it's possible PF here means Pred Forte and Durezol replaced it, or they are both being used. Given the encounter note lists PF 4/0 as current and Plan says continue Durezol QID, I'll include both as current OD medications. PFAT and gel drops are OTC (excluded per R2).", "task1_citation": "OD Citation: On Xal 0/1; Tim 0/1; PF 4/0 OS Citation: On Xal 0/1; Tim 0/1; PF 4/0", "task2_reasoning": "OD Reason: The extracted label includes both 'Pred Forte QID' and 'Durezol QID' for OD. Looking at the encounter note dated **/**/****, 'PF 4/0' is listed as a current medication, meaning Pred Forte QID OD. The Plan says 'Continue Durezol OD QID', which per R3 indicates Durezol is also current. However, it is clinically implausible that a patient would be on two different topical steroids (Pred Forte QID and Durezol QID) simultaneously in the same eye. More likely, 'PF 4/0' in the encounter notation refers to Pred Forte which was switched to or is the same as Durezol, or PF 4/0 actually IS Durezol (since the Plan explicitly says 'Continue Durezol OD QID'). The most reasonable interpretation is that PF 4/0 represents the steroid the patient is on, and the Plan clarifies that steroid is Durezol. Listing both Pred Forte QID and Durezol QID is likely a duplication error. The correct OD label should be 'Durezol QID' only (confirmed by Plan 'continue' trigger word). Citation: 'PF 4/0' and 'Continue Durezol OD QID' - these likely refer to the same medication, not two separate steroids. OS Reason: ", "task3_reasoning": "Priority 1: The current encounter (**/**/****) lists 'On Xal 0/1; Tim 0/1; PF 4/0'. Xal 0/1 = latanoprost QHS OS only. Tim 0/1 = timolol daily OS only. PF 4/0 = a steroid QID OD only. The Plan says 'Continue Durezol OD QID', which uses the trigger word 'continue' confirming Durezol is a current medication OD. It is clinically implausible that the patient is on both Pred Forte QID and Durezol QID in the same eye simultaneously. The 'PF 4/0' notation and 'Continue Durezol OD QID' most likely refer to the same steroid - Durezol. The Plan clarifies the identity of the steroid as Durezol. Therefore OD should list only Durezol QID, not both. PFATs (preservative-free artificial tears) are OTC and excluded per R2.", "task3_citation": "OD Citation: PF 4/0 ... Continue Durezol OD QID OS Citation: On Xal 0/1; Tim 0/1 ... Continue Timolol OS QAM and Latanoprost OS QHS"} |

| | | | | |
|---|---|---|---|---|
| CTM | Glaucoma type: POAG-severe OD, mild OS FH: mother, brother Tmax: 20/21 Pachymetry: OD 485(thin), OS 490(thin) Surgeries: CEIOL/STQ BGI 250 CPG AC 1 fen OD (*** **/**/****) Lasers: RT laser OD Medication failures/allergies: no kidney/breathing, Simbrinza - no effect Gonioscopy **/****: Gr 3, 3+pTM OU Comments: referred by Dr. *** (Trinidad), no renal or pulm issues, no h/o steroids No sleep apnea/migraines/hypotension Date IOP OD IOP OS Meds Notes **/**/**** 16 20/125- 15 Az 2/2, xalacom (xal/tim) qhs, Alpha 2/2 1st HVF, gonio, DFE, OCT **/**/**** 14 14 Az 2/2, alpha 2/2, Xalacom 1/1 IOP check **/**/**** 10 15 POD1 CE/BGI OD **/**/**** 13 16 PF 4/0, Moxi 4/0, Brom 1/0, Az 2/2, alpha 2/2, Xalacom 0/1 POW1 CE/BGI OD **/**/**** 15 17 PF 4/0, Ket 4/0, Az 2/2, Brim 2/2, Xalacom 0/1 POW 3 CE/BGI OD **/**/**** 28 11 PF 4/0, Moxi 4/0 , keto 4/0, Az 2/2, Agan 2/2, Xalacom 0/1 POW7 CE/BGI OD DFE, Mac OCT **/**/**** 16 16 Xalacom 0/1, Az 2/2, Alpha 2/2 POW10 CE/BGI OD **/**/**** 13 19 Xalacom 1/1, Az 2/2, Alpha 2/2 HVF, IOP **/**/**** 17 17 Az 2/2, Brim 3/3, Xalacom 1/1 qhs, not using tim brom 1/0 DFE, OCT, Mac OCT RNFL **/****: artifacts s/i thin L. 72 s/i thin RNFL **/****: R. 42 s/t/i thin, poor signal Mac OCT **/****: trace edema VF: **/**** superior depression, IAD MD -20 OD; full OS **/****: Diffuse depression, MD -21, L inferior deficits, MD -3.5 DFE: **/****, **/****: R. C/d 1.0, L. C/d 0.9 1. POAG-severe OD, moderate OS s/p CEIOL/CTR w STQ BGI 250 KPG AC 1 fen OD (*** **/**/****) - IOP acceptable OS, borderline OD - not on timolol - recommend tim qam/qam, xalacom qhs/qhs, continue Az 2/2, brim ok 3/3, continue brom 1/0 per Dr. *** - RSVP given tube - long discussion with patient regarding surgical outcome ... | brimonidine TID, brinzolamide BID, latanoprost/ timolol QHS | brimonidine TID, brinzolamide BID, bromfenac daily, latanoprost/ timolol QHS | {"task1_reasoning": "Priority 1: The current encounter dated **/**/**** lists medications: Az 2/2, Brim 3/3, Xalacom 1/1 qhs, and brom 1/0. 'not using tim' indicates timolol is not currently being used. Az 2/2 = Azopt BID OU. Brim 3/3 = brimonidine TID OU. Xalacom 1/1 qhs = Xalacom QHS OU (Xalacom is a combination product of latanoprost/timolol, kept as brand name). Brom 1/0 = bromfenac daily OD only (prescription non-glaucoma eyedrop for CME management, confirmed by Plan section 'continue brom 1/0 per Dr. ***'). The Plan section mentions 'recommend tim qam/qam' which is a new recommendation (future change, excluded) and 'continue Az 2/2, brim ok 3/3, continue brom 1/0' which confirms current use.", "task1_citation": "OD Citation: **/**/**** 17 17 Az 2/2, Brim 3/3, Xalacom 1/1 qhs, not using tim brom 1/0 OS Citation: **/**/**** 17 17 Az 2/2, Brim 3/3, Xalacom 1/1 qhs, not using tim", "task2_reasoning": "OD Reason: OS Reason: ", "task3_reasoning": null, "task3_citation": null} |

| | | | | |
|---|---|---|---|---|
| CTM | Impression Ms. *** is a 65 y.o. female POAG (reported) - follows with outside ophthalmologist - takes Agn 2/2 OU - ON appear healthy * Dry eye syndrome - With MGD - Scheduled preservative-free artificial tears every 2-4 hours - Refresh PM or other ointment at night - Lid hygiene with eyelids scrubs twice daily - Warm compresses twice daily for 10 minutes Blepharitis OU - Moderate MGD and scruf - Warm compresses 10 minutes twice a day - Lid hygiene with scrubs once daily<br>Today, **/**/****: · VA OD sc:20/40 cc: VA OS sc:20/50 cc: · IOP OD 22 / IOP OS 19 at 3:21 PM Applanation<br>.............................................................................. Disposition · Restart Brimonidine 2/2 · Continue AT's qid OU · Follow-up in 2 months TMax Goal CCT<br>Medications to avoid 552 541 **/**/**** Date VA OD VA OS IOP OD IOP OS<br>PG Beta CAI Cos Alpha Comb Simb Other G VF DFE OCT Pho Other | brimonidine BID | None | {"task1_reasoning": "Priority 1: The current encounter section (Impression) states the patient 'takes Agn 2/2 OU' (Alphagan/brimonidine BID OU). However, the Disposition/Plan section says 'Restart Brimonidine 2/2'. The word 'Restart' indicates the patient is NOT currently using brimonidine \u**** it is a future change (per R3, 'restart' = exclude). This creates a conflict with the Impression section which says 'takes Agn 2/2 OU'. The 'Restart' in the Plan strongly suggests the patient had stopped taking brimonidine and is being told to resume it. The Impression section likely reflects the historical/reported medication list rather than current active use. Since 'restart' implies the medication is not currently being used, brimonidine should be excluded. The medication table row for **/**/**** shows no entries under any medication column, confirming no current medications. Therefore, the output is None for both eyes.", "task1_citation": "OD Citation: takes Agn 2/2 OU OS Citation: takes Agn 2/2 OU", "task2_reasoning": "OD Reason: The note states 'takes Agn 2/2 OU' in the Impression section (Priority 1), which indicates the patient is currently taking Alphagan (brimonidine) BID in both eyes. While the Plan says 'Restart Brimonidine 2/2', the Priority 1 source explicitly states the patient 'takes' this medication. The Impression section describes the patient's current medication regimen as reported. The 'Restart' in the Plan could mean the patient had been inconsistent or had temporarily stopped, but the Priority 1 documentation says 'takes Agn 2/2 OU'. Per R4, if Priority 1 provides medication names, we should use Priority 1 and not override it with Plan section interpretation. The correct output should be 'brimonidine BID' for OD. OS Reason: Same reasoning as OD. The note states 'takes Agn 2/2 OU' in the Impression section (Priority 1), indicating current use of brimonidine BID in both eyes. The correct output should be 'brimonidine BID' for OS.", "task3_reasoning": "Priority 1: The Impression section dated to the current encounter (**/**/****) states 'takes Agn 2/2 OU', which indicates the patient is currently taking Alphagan (brimonidine) BID in both eyes. 'Agn' expands to Alphagan, which is brimonidine. The notation 2/2 means BID OD / BID OS. The Plan section says 'Restart Brimonidine 2/2', which could suggest the patient had been inconsistent, but the Priority 1 source explicitly uses the present tense 'takes', documenting current use. Per R4, Priority 1 takes precedence and we use the current encounter section. The Plan's 'Restart' is excluded per R3 (restart = future change, exclude). However, the Priority 1 documentation clearly states current use, so we extract from there.", "task3_citation": "OD Citation: takes Agn 2/2 OU OS Citation: takes Agn 2/2 OU"} |

| | | | | |
|---|---|---|---|---|
| ΔTM | Assessment: Pseudoexfoliation glaucoma OD Glaucoma suspect OS / PXS OS Presumed h/o narrow angles OU Reportedly diagnosed glaucoma ~**** in context of IOP elevation after multiple spinal steroid injections. Currently receiving eye care at *** with Dr. ***. Seen by glaucoma OD on **/**/****. S/p SLT OD sup hemisphere - **/**/**** S/p LPI OU -- patent OU S/p CE/IOL OU 2014 FHX none No h/o Trauma Current Flonase use Tmax 32/23 (ED visit **/**/****) CCT 505/507 Angles open OU (checked **/**/****) ONH cupping OU HVF 24-2 III **/****: OD generalized depression, possible sup arcuate; OS nonspecific defects RNFL/GCIPL **/****: OD insufficient signal; OS relatively full, ?early sup thinning AEs: H/o asthma/emphysema on home O2 -- timolol contraindicated; simbrinza caused redness/LL dermatitis; verticillata with rhopressa Sp SLT OD, minimal response to date IOP today persistently elevated OD Pseudophakia OU **** +Pseudophacodonesis OD > OS H/o recurrent subconj hemorrhage OU Plan: Latanoprost at bedtime both eyes Continue Dorzolamide 3 times daily both eyes Continue Methazolamide 25mg 2 times daily Continue Rhopressa at bedtime right eye Recommend establish PCP for routine medical care RBA AGI OD dw patient who agrees, signed consent, sent to preop, plan MAC-block AC tube | None | Start latanoprost QHS | {"task1_reasoning": "OD Citation and Reasoning: The Plan section states 'Latanoprost at bedtime both eyes'. Reviewing the current regimen earlier in the note, there is no mention of latanoprost being part of the current medication list. The note mentions dorzolamide, methazolamide (oral, excluded per R1), and Rhopressa as current medications. Latanoprost is a new addition. The Plan also states 'Continue Dorzolamide 3 times daily both eyes' and 'Continue Rhopressa at bedtime right eye', which are continuation statements (no change). Methazolamide is oral (excluded per R1). Per R6, latanoprost is a prostaglandin analogue, so frequency of once daily/at bedtime = QHS. OS Citation and Reasoning: The Plan section states 'Latanoprost at bedtime both eyes', which includes OS. Latanoprost is not part of the current regimen, so this is a new start. Per R6, prostaglandin frequency = QHS. The Plan also states 'Continue Dorzolamide 3 times daily both eyes', which is a continuation (no change). Rhopressa is only continued in OD. Methazolamide is oral and excluded per R1.", "task2_reasoning": "OD Reason: OS Reason: ", "task3_reasoning": null} |
| ΔTM | Summary: Diagnosed with glaucoma about 11 years ago. Previously followed by Dr. *** in MA. Has been followed by Dr. ***. - Outside glaucoma specialist: none - Surg hx: none - Laser hx: none - Glaucoma FHx: none - Tmax: unknown - CCT 557 / 548 - Gonio: open by Dr. *** (**/****) - Target IOP: ~mid teens OU - Med AEs: n/a **/**/****: IOP 13 / 14 On Lum 1/1 -- at target OU Imaging: HVF 24-2 (**/****) -- reliable, WNL - stable OD reliable, WNL - stable OS OCT RNFL (**/****) -- inf thinning - base OD inf thinning - base OS OCT GCIPL (**/****) -- WNL - base OD WNL - base OS ONH photo pending Assessment: 1. Primary open angle glaucoma mild OU IOP acceptable OU and OCT stable OU. 2. Nuclear sclerosis OU Not visually significant. Noticing some glare. 3. Dry Eye Syndrome OU Asymptomatic Plan: 1. Continue Lumigan OU QHS (okay to use every other day) 2. Monitor cataracts 3. ATs OU QID 4. MRx given Return in 6 months Tech: VA/MRx/PAP/glare/IOP Imaging: HVF | Decrease bimatoprost QOD | None | {"task1_reasoning": "OD Citation and Reasoning: The Plan section states 'Continue Lumigan OU QHS (okay to use every other day)'. The patient is currently on Lumigan 1/1 (QHS OU). The plan is to continue the same medication at the same frequency. The parenthetical '(okay to use every other day)' is permissive guidance, not a mandatory frequency change. ATs (artificial tears) are excluded per R1 as OTC. No changes documented for OD. OS Citation and Reasoning: The Plan section states 'Continue Lumigan OU QHS (okay to use every other day)'. The patient is currently on Lumigan 1/1 (QHS OU). The plan is to continue the same medication at the same frequency. The parenthetical '(okay to use every other day)' is permissive guidance, not a mandatory frequency change. ATs (artificial tears) are excluded per R1 as OTC. No changes documented for OS.", "task2_reasoning": "OD Reason: OS Reason: ", "task3_reasoning": null} |

| | | | | |
|---|---|---|---|---|
| ΔTM | Assessment: H/o bilateral uveitic glaucoma associated with uveitis with trabeculitis and very high IOP OU - H/o bilateral retinal hemes with temporal leakage on FA - Ddx possible Zika, dengue, chikungunya, rheumatologic. Bilateral HSV uncommon. - Zika negative in the serum and sperm sample - AC fluid virology for toxo, CMV, HSV, VZV, EBV **/**/**** ALL NEGATIVE - Quiescent OU today Uveitic glaucoma OU FH glaucoma (aunt) Gonio wide open Progressive cupping since **/**/****? Elevated IOP possible from inflammation vs. Steroid response Tmax 60 OU ** **** Cupping OS>OD s/p AGI OD **/**/**** s/p AGI OS **/**/**** HVF ** **** poor reliability with inferior arcuate OD, double arcuate OS; stable OU OCT RNFL/GCIPL November with diffuse thinning OU; appears worse compared to prior Progression on OCT compared to **/****, but also with period of IOP in high 20s to 30s from **/****-**/**** IOP today 13/14 on cosopt BID OU Plan: - Cosopt PF (blue cap) 2 times a day in both eyes - Artificial tears, restasis - *** 4 months, VA, IOP, dilate, OCTs - *** yearly | None | Start dorzolamide/ timolol PF BID, Stop dorzolamide/ timolol | {"task1_reasoning": "OD Citation and Reasoning: Current regimen shows 'IOP today 13/14 on cosopt BID OU'. Plan states 'Cosopt PF (blue cap) 2 times a day in both eyes'. Per R11, switching from traditional Cosopt to Cosopt PF (preservative-free) IS considered a change. This means stopping Cosopt and starting Cosopt PF BID for OD. OS Citation and Reasoning: Current regimen shows 'IOP today 13/14 on cosopt BID OU'. Plan states 'Cosopt PF (blue cap) 2 times a day in both eyes'. Per R11, switching from traditional Cosopt to Cosopt PF (preservative-free) IS considered a change. This means stopping Cosopt and starting Cosopt PF BID for OS.", "task2_reasoning": "OD Reason: OS Reason: ", "task3_reasoning": null} |
| ΔTM | Assessment: 1. s/p DSAEK OS (Dr. *** **/**/****) S/P PKP OS +pupilloplasty + sutured IOL, synechiolysis - h/o failed grafts x2 (high risk graft) **/**/**** - h/o trauma OD - Graft attached and clear - Abnormal blood vessels, will go away (**/**/****) 2. Pseudophakia OD **/**/**** Topo suspicious for KCN 3. Traumatic Glaucoma vs. CACG OS H/o childhood trauma at age 6 Sp PK x 3 Per pt, IOP well controlled on cosopt and alphagan Cupping OS>OD OCT RNFL/GCIPL ** **** full OD, thinned OS, worse OS compared to **** HVF ** **** full OD, stable; sim V constricted OS, worse compared to **** IOP stable on Cosopt 0/2, Alphagan 0/2 LTFU from **** to **** Plan: Continue cosopt 3 times a day left eye Discontinue alphagan left eye (redness, irritation) Start latanoprost at bedtime left eye Predforte 3 times a day left eye RTC 4-6 months *** and ***, IOP check, consider Yag OS w Dr. *** - IOP check in Aruba in 1 week, adjust medical therapy as needed to keep IOP around 15 mm Hg | Start latanoprost QHS, Stop brimonidine, Increase dorzolamide/ timolol TID | Start latanoprost QHS, Start prednisolone TID, Stop brimonidine, Increase dorzolamide/ timolol TID | {"task1_reasoning": "OD Citation and Reasoning: No changes documented for the right eye. Current medications are Cosopt 0/2 and Alphagan 0/2, meaning only OS is being treated. The Plan section documents changes only for OS. OS Citation and Reasoning: Current regimen: Cosopt 0/2 (BID OS), Alphagan 0/2 (BID OS). Plan states: 'Continue cosopt 3 times a day left eye' - Cosopt was BID (0/2) and is now TID for OS, so this is an increase. 'Discontinue alphagan left eye' - stopping Alphagan OS. 'Start latanoprost at bedtime left eye' - new prostaglandin, per R6 labeled QHS. 'Predforte 3 times a day left eye' - Pred Forte not in current regimen, so this is a new start at TID.", "task2_reasoning": "OD Reason: OS Reason: ", "task3_reasoning": null} |

| | | | | |
|---|---|---|---|---|
| COM | Impressions 1. Primary Open Angle Glaucoma OD >> OS, diagnosed in **** Normal CCT, Tmax 27 per patient HVF with central island OD, OS with inferior arcuate and superior nasal step which appears progressed from **** Repeat today (**/**/****): generalized depression OD on 10-2, worsening IA on 24-2 OS Refused HVF on **/**/**** Baseline OCT RNFL (**/**/****) with diffuse thinning OU IOP improved to mid-to-upper teens with neptazane BID (tolerating well) but not at goal for OD; compliance significantly improved At visit on **/**** he said he ran out of xalatan 3 months ago (although he has a refill from last visit for 1 year) Was previously using brimonidine 1/1 (says he ran out because not enough in bottle although Rx written for TID) Sensory XT OD 2. S/p LPI OS (Dr. *** in ****) 3. Non-visually significant cataracts OU Plan: 1. Continue Cosopt BID OU Xalatan hs/hs Brimonidine to 2/2 Neptazane 50 mg PO TID; patient cannot take TID dosing due to work so recommend taking 2 tabs in AM and one in PM 2. Again had extensive discussion regarding the need for surgery OD before OS especially in light of the fact that he has been noncompliant with meds, IOP is elevated, and he has advanced field loss with evidence of progression OS. Surgery has been recommended since **/****. I have recommended surgery OD before OS but patient is hesitant and is concerned that he will miss work (carpet cleaner) after surgery. Patient was seen last visit with Dr. ***, who also recommended urgent surgery. He understands that if IOP is not better controlled, he will lose more vision, and that vision loss is permanent. Today, patient reports that he is having difficulty from work and is worried about losing job if taking too much time off, but he understands that IOP needs to be lower. I have written him a note to his work explaining situation. Patient will try increasing PO meds and discuss<br>surgery with his wife and job. If he still refuses surgery, SLT could be tried to offer some minimal IOP lowering effect 4. RTC 1-2 mo IOP check and surgery decision TMax Goal CCT Medications to avoid 27 532/534 Date VA OD VA OS IOP OD IOP OS PG Beta CAI Cos Alpha Comb Simb Other G VF DFE OCT Pho Other **/**/**** 70 25 19 15 x x x x x x **/**/**** 200 40 13 14 x x x x x x **/**/**** 200 20 24 22 2/2 1/1 x **/**/**** 200 25 16 17 1/1 2/2 2/2 Neptazane 50 BID Increase neptazane to 50 TID **/**/**** 200 25 19 17 1/1 2/2 2/2 Neptazane 50 BID **/**/**** 200 25 17 14 1/1 2/2 2/2 Neptazane 50 BID Refused HVF today | methazolamide 50mg BID | methazolamide 50mg TID | {"task1_reasoning": "Oral Citation and Reasoning: The Plan section states: 'Continue... Neptazane 50 mg PO TID; patient cannot take TID dosing due to work so recommend taking 2 tabs in AM and one in PM.' The word 'Continue' explicitly confirms current use per R2 (Priority 2). Neptazane is the brand name for methazolamide. Dose is 50mg, frequency is TID. Per R3, route (PO) and extended release info are excluded from output.", "task2_reasoning": "Oral Reason: ", "task3_reasoning": null} |

| | | | | |
|---|---|---|---|---|
| ΔOM | **/**/****: Able to review paper chart dating back to **** Assessment: 1. Advanced OAG, Both Eyes OS>>OD Patient had a Trabeculectomy **/**/**** by Dr. *** with Mitomycin but had postop IOP spike and believes lost fixation / Wipe out of central Va 1st day after surgery Patient had vision of 20/50 OS **/**/**** with extensive VF loss Has never had laser in either eye Patient is VERY resistant to future surgery Patient tried on Neptazane in **** but was only on 25 BID. Had some effect OD, not OS. Pt believes he tolerated medication Current Regimen Travatan Z hs / hs Cosopt 2 / 2 Alphagan 3 / 3 Methazolamide 50mg po BID (was started in March ****, patient ran out after one month and did not have any refills, per patient) **/**/**** HVF OD baseline for 24-2 size V, superior arcuate near fixation and inferior paracentral arcuate, OS size V 24-2 poorly reliable with generalized depression, least inferiorly IOP better at 10 / 10 today (**/**/****). Patient refused Esterman visual field today. Does not want to do any more VFs. Thin CCT 2. Early Glare / Early Cataract, OU Recommend Amber glass 3. Failed Trab, OS This could be working a little as there is a thick bleb Plan: 1. Continue present management. Continue The MZM / reordered. 2. Monitor cataracts. 3. Patient drives: Needs Esterman field (refused today). 5. Long discussion with patient about risk & benefit of surgery. 5. Return in 3 months for IOP check, Esterman HVF and dilation. | Start methazolamide 50mg BID | None | {"task1_reasoning": "Oral Citation and Reasoning: The Plan section for the encounter date **/**/**** states: 'Continue present management. Continue The MZM / reordered.' This indicates continuation of the current methazolamide regimen (50mg BID as noted in the current regimen). The MZM was reordered (refilled), but no change in dose or frequency was made. Per R9, continuation statements indicate no changes.", "task2_reasoning": "Oral Reason: ", "task3_reasoning": null} |

*CTM = current topical medications, COM = current oral medications, ΔTM = change in topical medications, ΔOM = change in oral medications*

*Any names, dates, or other potentially identifying information have been replaced by asterisks.*
